# COVID MED – An Early Pandemic Randomized Clinical Trial of Losartan for Hospitalized COVID-19 Patients

**DOI:** 10.1101/2022.01.12.22269095

**Authors:** Daniel Freilich, Jennifer Victory, Paul Jenkins, James Wheeler, G. Matthew Vail, Erik Riesenfeld, Peggy Cross, Catherine Gilmore, Melissa Huckabone, Anna Schworm, Umesha Boregowda, Farah Deshmukh, Yuri Choi, Azkia Khan, Anne Gadomski

## Abstract

**Background:** ACEi/ARB medications have been hypothesized to have potential benefit in COVID-19. Despite concern for increased ACE-2 expression in some animal models, preclinical and observational-retrospective and uncontrolled trials suggested possible benefit. Two RCTs of the ARB losartan from University of Minnesota showed no benefit yet safety signals for losartan in outpatient and hospitalized COVID-19 patients. COVID MED, started early in the pandemic, also assessed losartan in a RCT in hospitalized patients with COVID-19.

**Methods:** COVID MED was a double-blinded, placebo-controlled, multicenter, platform randomized clinical trial (RCT). Hospitalized COVID-19 patients were randomized to receive standard care and hydroxychloroquine, lopinavir/ritonavir, losartan, or placebo. Hydroxychloroquine and lopinavir/ritonavir arms were discontinued after RCTs showed no benefit. We report data from the losartan arm compared to combined (lopinavir-ritonavir and placebo) and prespecified placebo-only controls. The primary endpoint, the mean COVID-19 Ordinal Severity Score (COSS) slope of change, was compared with the Student’s t-test. Slow enrollment prompted early termination.

**Results:** Of 448 screened patients, 15 (3.5%) were randomized/enrolled, 9 to receive losartan and 6 to receive control (lopinavir/ritonavir [N=2], placebo [N=4]); 1 patient who withdrew prior to study drug was excluded yielding 14 patients for analysis (losartan [N=9] vs. control [N=5] [lopinavir/ritonavir [N=2], placebo [N=3]]). Most baseline parameters were balanced. Treatment with losartan was not associated with a difference in mean COSS slope of change in comparison with combined control (p=0.4) or placebo-only control (p=0.05) (trend favoring placebo). 60-day mortality and overall AE and SAE rates were numerically but not significantly higher with losartan.

**Conclusions:** In this small blinded RCT in hospitalized COVID-19 patients, losartan did not improve outcome vs. control comparisons and was associated with adverse safety signals.

## INTRODUCTION

Angiotensin converting enzyme inhibitors (ACEi) and angiotensin II receptor blockers (ARB) have been hypothesized to have benefit in COVID-19 secondary to inhibition of pneumocyte ACE-2 receptors resulting in decreased SARS-CoV-2 entry, as well as local vasodilation/vasoconstriction alteration and anti-inflammatory effects. Animal studies suggested potential for benefit. Safety concerns have included observations of increased ACE-2 receptor expression in some animal models and potential for medication class adverse events (e.g., acute kidney injury [AKI]). Retrospective and observational continuation-discontinuation trials [1-10], open-label interventional trials [11-15], and a meta-analysis [16] showed trends favoring benefit. Two blinded RCTs, however, showed no benefit yet adverse safety signals [17-18]. We initiated the COVID MED platform RCT at the pandemic onset prior to publication of ACEi/ARB COVID-19 clinical trials, aiming to assess the ARB losartan (and hydroxychloroquine and lopinavir/ritonavir) in hospitalized COVID-19 patients.

## MATERIALS AND METHODS

COVID MED (NCT04340557), a parallel design blinded platform RCT, was approved by the Institutional Review Boards of Bassett Medical Center (Cooperstown, NY [April 3, 2020] [#1581969]), Goshen Health (Goshen, IN) and Reid Health (Richmond, IN). Initiated at the pandemic onset, we could not project COVID-19 admission numbers, thus, an arbitrary N of 4,000 hospitalized patients with laboratory-confirmed SARS-CoV-2 (screening criteria) was selected with prespecification for sample size/power calculations and updating after interim analysis. Our primary objective was to assess whether treatment with hydroxychloroquine, lopinavir/ritonavir, or losartan, would result in more rapid improvement in the 7-point COVID-19 Ordinal Scale Score (COSS) in comparison with placebo. COSS is as follows: 1) Death; 2) Hospitalized, on mechanical ventilation or ECMO; 3) Hospitalized, on NIV/high flow oxygen; 4) Hospitalized, requiring oxygen; 5) Hospitalized, not requiring oxygen; 6) Not hospitalized, with limitations; 7) Not hospitalized, without limitations. Enrolled patients were followed for up to 60 days.

Target recruitment was maximized by daily assessment of positive SARS-CoV-2 swab tests from Bassett’s laboratory in hospitalized patients with review of inclusion/exclusion criteria for all such patients by the study’s enrollment nurse and offering enrollment to all interested in participating after a comprehensive informed consent process (see Supplementary Material for Informed Consent Form [ICF] and Protocol). Post-consent, patients were randomly assigned by an unblinded enrollment research nurse in a 2:2:2:1 ratio in blocks to one of four groups using a computer-generated randomization schedule provided by the study’s statistician (without stratification): hydroxychloroquine, lopinavir/ritonavir, losartan, or placebo; this ratio was selected because early in the pandemic patients declined participation in trials with low likelihood of receiving ‘active’ drug. Allocation concealment was ensured by having only the enrollment research nurse be unblinded and with maintaining confidentiality of the allocation.

All groups received standard care, which transformed over time. Hydroxychloroquine and lopinavir/ritonavir enrollment were halted after other RCTs showed no benefit [19-20]; a 2:1 ratio computer-generated randomization schedule for losartan and placebo was used thereafter. Herein, we focus on the trial design and results for losartan vs. combined control groups (we included 2 lopinavir/ritonavir patients as other RCTs showed no benefit or safety concerns [20]. We also report outcome data for the comparison of the losartan and placebo-only group, per our prespecified statistical analysis plan (SAP). Low enrollment led to study termination May 27, 2021.

Inclusion criteria were (1) hospitalized; (2) >18 years-old; (3) laboratory-confirmed SARS-CoV-2 infection; (4) randomization within 72 hours of admission; and (5) able to provide consent. General exclusion criteria applicable to all groups included (1) ESRD; (2) severe hepatic insufficiency; (3) nausea/vomiting/aspiration risk precluding oral medications unless can be given by nasogastric tube; (4) use of another SARS-CoV-2 medication; (5) pregnancy/breast feeding; (6) no contraception; and (7) inability to obtain consent. Each treatment group had specific exclusions based on the medication’s safety profile. Specific losartan group exclusion criteria included (1) ARB allergy/intolerance; (2) taking ACEi/ARB; (3) hypotension; (4) hyperkalemia; (4) severe renal dysfunction; (5) severe volume depletion/AKI; (6) ascites; (7) aortic/mitral stenosis; (8) renal artery stenosis; and (9) co-administration of CYP3A interacting drugs. Minor modifications were made during the study. There were no placebo group specific exclusion criteria. After hydroxychloroquine and lopinavir-ritonavir enrollment was stopped, inclusion/exclusion criteria were the same for the losartan and placebo groups.

Study drug (losartan or TicTacs placebo [lopinavir-ritonavir in 2 patients]) was prepared by a pharmacist who inserted the ‘pills’ into blank capsules – enabling cost-effective quadruple-blinding (patient, clinical nurse/physician, study nurse, investigators); only the enrollment study nurse was unblinded but he/she had no other responsibilities after enrollment. Unblinding was not done. If crushed drug or drug in solution had to be administered (nasogastric/gastric tube), eye shields were used to maintain blinding. Dosing was initially twice daily in all groups to mask varying regimens (losartan group: losartan and placebo; placebo group: placebo and placebo; lopinavir/ritonavir group: lopinavir/ritonavir twice daily). The losartan dose was 25 mg; the lopinavir/ritonavir dose was 400/100 mg. After hydroxychloroquine and lopinavir/ritonavir groups enrollment ceased, losartan and placebo dosing were daily. Dosing duration target was 14 days (minimum 5 days) if tolerated (no clinically relevant related AEs/SAEs such as hypotension, hyperkalemia, or worsening renal function). Adherence to study dosing was monitored and noted by the study’s research nurse. Patients discharged early were administered a shorter 5-day course with a minimum of 1.5 days as an in-patient (3 doses) and 3.5 days as an out-patient (7 doses).

With the initial study plan to recruit up to 4,000 subjects, the 2:2:2:1 assignment was predicted to result in 571 subjects in the placebo group and 1,143 subjects in each treatment group. An independent, conflict-free, and multidisciplinary Data Monitoring Committee (DMC) was convened at the study’s outset and agreed to its processes and goals; unexpected SAE reports were shared with the DMC. Interim analysis was planned for each treatment group at an information fraction of 0.1, which would occur when a treatment group had slopes for 114 subjects, and the control group has slopes for 57. After hydroxychloroquine and lopinavir/ritonavir were dropped, we planned interim analysis and sample size optimization upon enrollment of 114 losartan and 57 placebo subjects. However, the study was stopped due to slow enrollment prior to the first planned DMC interim analysis.

Protocol CRFs were used to collect standardized efficacy/safety data; telephone follow-up was used as much as possible after discharge to obtain planned efficacy/safety data but inability to meet participants due to COVID-19 safety restrictions precluded complete data collection. Additional blood tests for more extensive safety monitoring were completed for patients who consented. AEs were classified in accordance with NCICTC for Adverse Events, version 4.0 (standard grading procedures). COVID MED was carried out in accordance with principles of the Code of Ethics of the World Medical Association (Declaration of Helsinki) and GCP guidelines of the International Conference on Harmonization, with general principles of protection of humans participating in research. Data from CRFs and AE/SAE reports were entered into REDCap using secure login/password entry; data were deidentified in working documents which were available only to study investigators; all study identifying data will be destroyed/deleted upon publication to allow wider sharing if requested.

### Statistical Analysis

As per our prespecified SAP, subjects had COSS values recorded for up to 11 timepoints (days 0, 1-7, 14, 30, and 60). These 11 X (time) Y (COSS) pairs were be used to calculate the mean slope of the change in COSS values over time for each subject. Only X-Y pairs up to and including the first attainment of COSS level 7 were included. These mean slopes of the change in COSS levels over time served as the primary endpoint for each subject.

The study design was such that it constituted three unique trials, all of which utilized the same control (placebo) group. Means of the slopes for each of the three different treatments were compared separately to mean slope in the control group using Student’s t-test (rather than Kaplan-Meier assessment as planned in the SAP). We report methods/results for comparison of the losartan vs. placebo-only arms per the prespecified SAP, but also, because of our small N, post-hoc comparison with combined controls (lopinavir/ritonavir and placebo). One important additional deviation from the SAP was completion/reporting herein of per protocol (a planned secondary outcome) rather than intention-to-treat (the planned primary outcome) analysis, also because of the final small N (this change was decided on prior to review of individual patient outcome data).

Continuous and categorical secondary endpoints were compared using Student’s t-test and Fisher’s exact test, respectively. Secondary analyses included but were not limited to hospital LOS, mechanical ventilation duration, and 60-day mortality. Time to resolution of nasopharyngeal viral shedding (PCR) was compared. For safety, the primary outcome was the 60-day overall SAE rate; secondary measurements included overall AE rates, and AKI, hypotension, and hyperkalemia AE/SAE rates. Adjusted and subgroups analyses were not done due to small sample size. For the primary outcome, missing values were addressed with conventional last-observation-carried-forward (LOCF).

## RESULTS

### Screening/enrollment

Of 448 screened patients, 15 were consented, randomized, and enrolled (3.3%), 9 receiving losartan and 6 receiving control (placebo=4, lopinavir/ritonavir=2); 1 placebo patient who withdrew after enrollment but prior to study drug administration was excluded (yielding 5 control patients [placebo=3, lopinavir/ritonavir=2] for analysis). Reasons for non-inclusion at Bassett: patient declined (12.0%), taking ACEi/ARB (26.8%), outside enrollment window (20.1%), hypotension (1.6%), hyperkalemia (2.1%), renal disease (4.2%), altered mental status precluding consent (11.5%), hospitalized for non-COVID-19 reasons (asymptomatic) (10.4%), withdrawal post-consenting but pre-randomization (0.2%), and other (11.1%) (see Figure 1).

**Figure 1.**
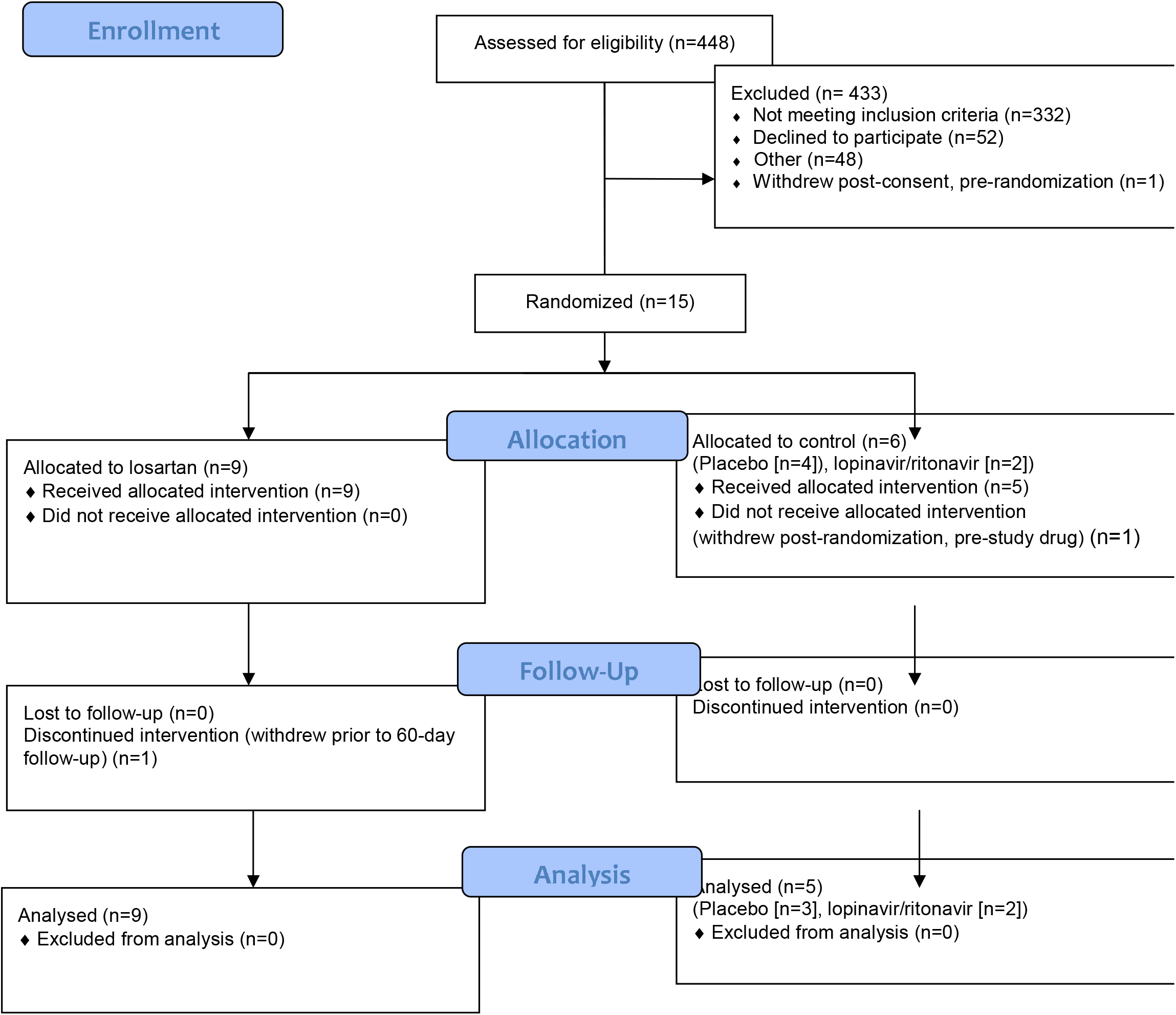
CONSORT 2010 Flow Diagram (http://www.consort-statement.org/)

### Baseline parameters/demographics

Comparisons of losartan and combined control baseline data revealed balanced parameters other than comorbidities. Specifically, mean age was 63.7 vs. 61.8 years, and males accounted for 66.7% vs. 60%, respectively. 100% were Caucasian and 100% were ward patients in both groups. Mean COSS was 3.6 vs. 4.0, (p=0.5), qSOFA 0.33 and 0.6 (p=0.4), and creatinine 0.7 vs. 0.9 (p=0.4), respectively. Chest x-ray opacities consistent with pneumonia were seen in 88.9 vs. 100% (p=0.9). PSI scores were similar – 54.9 and 59.3 (p=0.5). Mean BMI was 31.0 and 31.8 (p=0.4). The mean targeted comorbidities rate (per subject) was lower with losartan than combined control (1 vs. 2.6, p=0.02); a similar trend was seen for the Charlson Score (2.5 vs. 4.5, p=0.08). Treatment occurred at a median of 9.3 vs. 7.4 days after symptoms onset (p=0.3). Treatment duration mean was 9.6 vs. 13.3 days, respectively (p=0.3). 88.9 vs. 50% of patients received corticosteroids (p=0.5). Most patients were enrolled between the spring of 2020 and the winter of 2021 (88.9 vs. 80%). Baseline comparisons for losartan vs. placebo-only control were similar (see Table 1).

**Table 1.** Baseline parameters/demographics. Losartan is compared with combined control (lopinavir/ritonavir and placebo) and placebo only control. Statistically significant and ‘trend’ comparisons are in bold.

|  | Treatment | Combined control | p | 95% CI | Placebo only control | p | 95% CI |
| --- | --- | --- | --- | --- | --- | --- | --- |
|  | Losartan | Lopinavir/ritonavir and placebo |  |  | Placebo |  |  |
| N | 9 | 5 | NA | NA | 3 | NA | NA |
| Age (mean) | 63.7 | 61.8 | 0.8 | -17.0153 to 13.2153 | 56.3 | 0.4 | -27.4633 to 12.6633 |
| Male (%) | 66.7 | 60.0 | 0.9 | -0.80986 to 0.94319 | 66.7 | NA | NA |
| Enrollment spring 2020 | 11.1 | 60 | 0.1 | -1.0732 to 0.0955 | 33.3 | 0.4 | -0.7557 to 0.3112 |
| Enrollment fall 2020 - winter 2021 | 77.8 | 20 | 0.2 | -0.2486 to 1.4042 | 33.3 | 0.4 | -0.6224 to 1.5113 |
| Enrollment spring 2021 | 11.1 | 20 | 0.7 | -0.50209 to 0.32431 | 33.3 | 0.4 | -0.7557 to 0.3112 |
| Caucasian ethnicity (%) | 100 | 100 | NA | NA | 100 | NA | NA |
| COSS (mean) | 3.6 | 4.0 | 0.5 | -0.7192 to 1.5192 | 3.7 | 0.9 | -1.2961 to 1.4961 |
| Symptoms duration, days (mean) | 9.3 | 7.4 | 0.3 | -6.0157 to 2.315 | 6.7 | 0.3 | -7.9688 to 2.8688 |
| Comorbidities (targeted) rate (mean) | 1.0 | 2.6 | **0.02** | 0.3255 to 2.8745 | 2.7 | **0.046** | 0.0392 to 3.3608 |
| Immunocompromised (%) | 22.2 | 60 | 0.3 | -1.0311 to 0.2755 | 66.7 | 0.2 | -1.1988 to 0.3099 |
| Chronic heart disease (%) | 22.2 | 80 | 0.1 | -1.2935 to 0.1379 | 66.7 | 0.2 | -1.1988 to 0.3099 |
| Chronic lung disease (%) | 33.3 | 60 | 0.5 | -0.9823 to 0.449 | 66.7 | 0.5 | -1.1768 to 0.5101 |
| Chronic kidney disease (%) | 0 | 0 | NA | NA | 0 | NA | NA |
| Chronic liver disease (%) | 0 | 0 | NA | NA | 0 | NA | NA |
| Diabetes (%) | 22.2 | 25 | 0.9 | -0.48384 to 0.52828 | 33.3 | 0.7 | -0.7644 to 0.5422 |
| Extreme obesity (%) | 0 | 50 | **0.03** | -0.962 to -0.038 | 50 | **0.03** | -0.962 to -0.038 |
| Charlson Score (mean) | 2.5 | 4.5 | **0.08** | -0.3196 to 4.3196 | 3.7 | 0.3 | -1.1366 to 3.5366 |
| qSOFA (mean) | 0.33 | 0.6 | 0.4 | -0.3585 to 0.8985 | 0.3 | 1 | -0.7680 to 0.7680 |
| BMI (mean) | 31.0 | 31.8 | 0.4 | -4.2764 to 9.2764 | 35.1 | 0.2 | -2.3076 to 10.5076 |
| Creatinine (mean) | 0.7 | 0.9 | 0.4 | -0.2508 to 0.5708 | 0.7 | 0.9 | -0.2127 to 0.2327 |
| Chest x-ray opacities (%) | 88.9 | 100 | 0.9 | -1.1646 to 0.9423 | 100 | 0.9 | -1.3621 to 1.1399 |
| Pneumonia severity index (PSI) (mean) | 54.9 | 59.3 | 0.5 | -41.3071 to 50.1071 | 60 | 0.8 | -40.4537 to 21.2537 |
| Treatment days (mean) | 9.6 | 13.3 | 0.3 | -3.5545 to 10.8545 | 13 | 0.4 | -5.1086 to 11.9086 |
| Treatment with corticosteroids (%) | 88.9 | 40 | 0.3 | -0.7949 to 0.2172 | 60 | 0.7 | -0.9706 to 1.415 |

### Efficacy

COSS data were recorded for 97% of losartan group timepoints (3% missingness), 94.5% of combined control (lopinavir/ritonavir and placebo) group timepoints, and 100% of placebo-only control group timepoints. The mean slopes of the change for COSS were 0.00365 for losartan (p=0.5), 0.02091 for combined control (p=0.07), and 0.05268 for placebo-only control (p=0.002). Comparisons of the slopes of the change for COSS, our primary outcome measurement, revealed: losartan vs. combined control (p=0.4), losartan vs. placebo-only control (p=0.05) (trend favoring placebo), combined control vs. placebo-only control (p=0.267) (see Figure 2).

**Figure 2.**
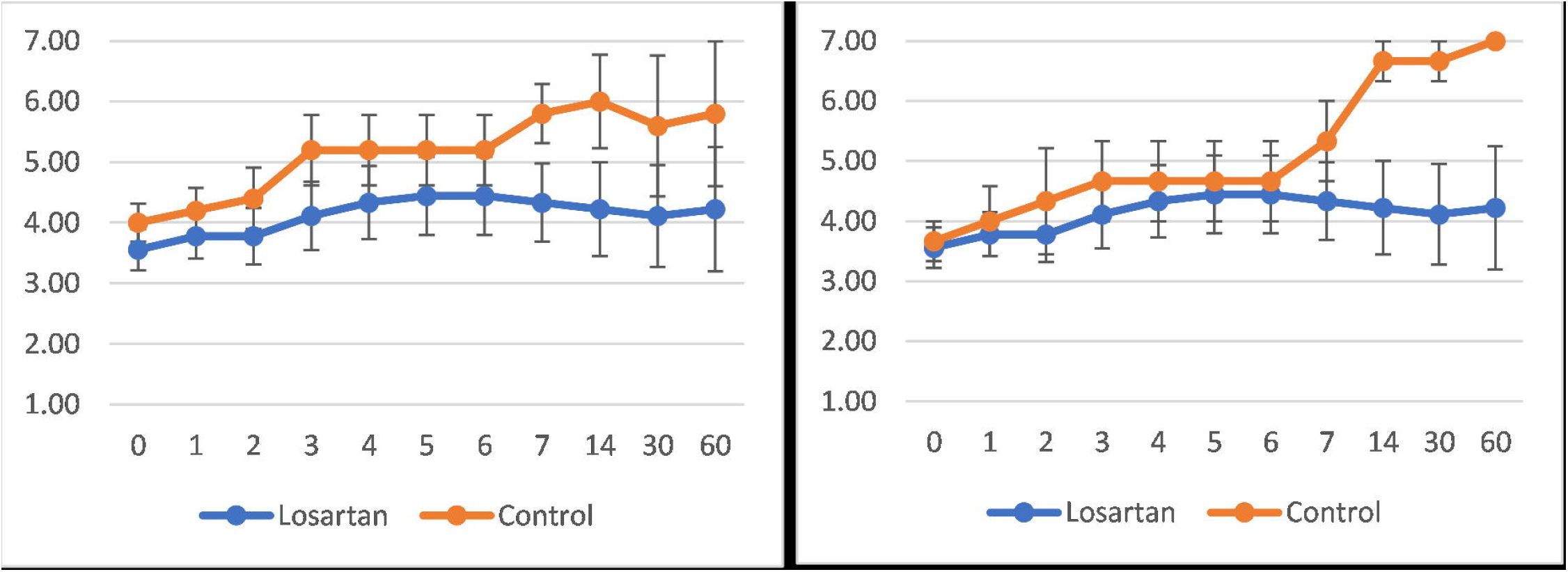
COSS over time +/- SEM. Figure on left compares losartan and combined control (lopinavir/ritonavir and placebo). Figure on right compares losartan and placebo only control. Comparisons of the mean of the slopes of the change for COSS, our primary outcome, revealed: losartan vs. combined control (p=0.4), losartan vs. placebo only control (p=0.05) (trend favoring placebo), combined control vs. placebo only control (p=0.267).

Mean COSS change at 60 days was numerically but not significantly lower with losartan (0.7) than combined control (1.8) (p=0.6). Mortality at 60 days (44.4 vs. 20%, p=0.9), mean LOS (16.4 vs. 7.0 days, p=0.2), and mean mechanical ventilation duration (8.5 vs. 0 days) were worse with losartan but also not significantly (see Table 2).

**Table 2.**
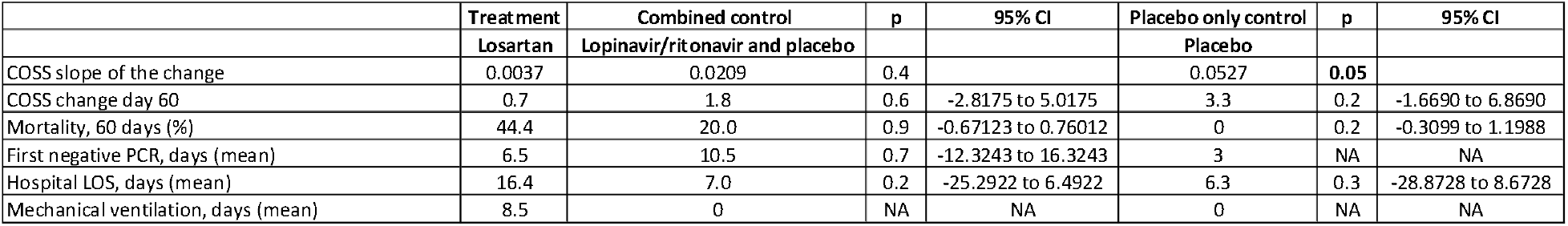
Efficacy. Losartan is compared with combined control (lopinavir/ritonavir and placebo) and placebo only control. Statistically significant and ‘trend’ comparisons are in bold. Comparison of mean COSS slope of the change was the study’s primary efficacy outcome measurement.

|  | Treatment | Combined control | p | 95% CI | Placebo only control | p | 95% CI |
| --- | --- | --- | --- | --- | --- | --- | --- |
|  | Losartan | Lopinavir/ritonavir and placebo |  |  | Placebo |  |  |
| COSs slope of the change | 0.0037 | 0.0209 | 0.4 |  | 0.0527 | **0.05** |  |
| COSs change day 60 | 0.7 | 1.8 | 0.6 | -2.8175 to 5.0175 | 3.3 | 0.2 | -1.6690 to 6.8690 |
| Mortality, 60 days (%) | 44.4 | 20.0 | 0.9 | -0.67123 to 0.76012 | 0 | 0.2 | -0.3099 to 1.1988 |
| First negative PCR, days (mean) | 6.5 | 10.5 | 0.7 | -12.3243 to 16.3243 | 3 | NA | NA |
| Hospital LOS, days (mean) | 16.4 | 7.0 | 0.2 | -25.2922 to 6.4922 | 6.3 | 0.3 | -28.8728 to 8.6728 |
| Mechanical ventilation, days (mean) | 8.5 | 0 | NA | NA | 0 | NA | NA |

### Safety

The primary safety outcome measurement, the overall SAE rate (per subject), was numerically higher with losartan vs. combined control (2.0 vs. 0.6%) and vs. the placebo-only control (2.0 vs. 0%). The overall AE rate (per subject) trend was similar, being higher with losartan than combined control (3.9 vs. 1.0%) and vs. placebo-only control (3.9 vs. 0%). AKI AE and SAE rates were similar, but hypotension and hyperkalemia, as well as respiratory failure AE and SAE rates, were numerically higher with losartan than combined control and placebo-only control. No safety comparison was statistically significantly different (see Table 3).

**Table 3.** Safety. AE and SAE rates including relatedness. Losartan is compared with combined control (lopinavir/ritonavir and placebo) and placebo only control. Statistically significant and ‘trend’ comparisons are in bold. Comparison of SAE rate was the study’s primary safety outcome measurement.

|  | Losartan |  |  |  |  | Control (placebo or lopinavir/ritonavir) |  |  |  |  | p | 95% CI | Control (placebo) |  |  |  |  | p | 95% CI |
| --- | --- | --- | --- | --- | --- | --- | --- | --- | --- | --- | --- | --- | --- | --- | --- | --- | --- | --- | --- |
|  | Total | Definitely | Probably | Possibly | Not | Total | Definitely | Probably | Possibly | Not |  |  | Total | Definitely | Probably | Possibly | Not |  |  |
| AEs/subject (mean) | 3.9 | 0 | 0 | 1.0 | 2.9 | 1.0 | 0 | 0 | 1.0 | 0 | 0.3 | -8.3414 to 2.5414 | 0 | 0 | 0 | 0 | 0 | NA | NA |
| SAEs/subject (mean) | 2.0 | 0 | 0 | 0.4 | 1.6 | 0.60 | 0 | 0 | 0.6 | 0 | 0.3 | -4.3144 to 1.5144 | 0 | 0 | 0 | 0 | 0 | NA | NA |
| AKI AEs/subject (mean) | 0.22 | 0 | 0 | 0.11 | 0.11 | 0.20 | 0 | 0 | 0.2 | 0 | 0.95 | -0.7560 to 0.7160 | 0 | 0 | 0 | 0 | 0 | NA | NA |
| AKI SAEs/subject (mean) | 0.11 | 0 | 0 | 0.00 | 0.11 | 0.20 | 0 | 0 | 0.2 | 0 | 0.5 | -0.5160 to 0.9560 | 0 | 0 | 0 | 0 | 0 | NA | NA |
| Hypotension AEs/subject (mean) | 0.33 | 0 | 0 | 0.22 | 0.11 | 0 | 0 | 0 | 0 | 0 | NA | NA | 0 | 0 | 0 | 0 | 0 | NA | NA |
| Hypotension SAEs/subject (mean) | 0.22 | 0 | 0 | 0.11 | 0.11 | 0 | 0 | 0 | 0 | 0 | NA | NA | 0 | 0 | 0 | 0 | 0 | NA | NA |
| Hyperkalemia AEs/subject (mean) | 0.11 | 0 | 0 | 0.11 | 0 | 0 | 0 | 0 | 0 | 0 | NA | NA | 0 | 0 | 0 | 0 | 0 | NA | NA |
| Hyperkalemia SAEs/subject (mean) | 0.00 | 0 | 0 | 0 | 0 | 0 | 0 | 0 | 0 | 0 | NA | NA | 0 | 0 | 0 | 0 | 0 | NA | NA |
| Respiratory failure/COVID-19 PNA AEs rate (mean) | 0.67 | 0 | 0 | 0.33 | 0.33 | 0.20 | 0 | 0 | 0.2 | 0 | 0.2 | -1.2420 to 0.3020 | 0 | 0 | 0 | 0 | 0 | NA | NA |
| Respiratory failure/COVID-19 PNA SAEs rate (mean) | 0.67 | 0 | 0 | 0.33 | 0.33 | 0.20 | 0 | 0 | 0.2 | 0 | 0.2 | -1.2420 to 0.3020 | 0 | 0 | 0 | 0 | 0 | NA | NA |

Planned subgroups analysis and adjustment for baseline parameters was not done due to the small N of the enrolled population.

## DISCUSSION

COVID MED, to our knowledge, the third blinded, placebo-controlled RCT assessing an ARB in COVID-19, did not find significant group differences in the COSS score for hospitalized patients treated with losartan vs. control. A statistically insignificant trend favoring control was found for our prespecified primary efficacy outcome measurement, comparison of the mean COSS slope of the change vs. placebo-only control (p=0.05). Our primary safety outcome measurement, overall SAE rate, was numerically but not statistically significantly higher with losartan. Secondary outcomes also numerically favored placebo but no group comparisons were statistically significantly different – including mortality.

Although COVID MED was small and pilot-like in scope, its results are value-added in that they add randomized, blinded, and placebo-controlled data to the limited ACEi/ARB literature in COVID-19. Our results are similar to those found in two larger blinded RCTs [17-18]; these three RCTs do not support empiric starting of ACEi/ARB, at least ARB, in COVID-19 outside of RCTs. Given the negative results of these blinded studies, it is worth reviewing the evidence for ACEi/ARB in COVID-19.

### Preclinical data

SARS-CoV and SARS-CoV-2 viruses enter cells by binding to cellular ACE2 receptors which are expressed on lung type II pneumocytes, enterocytes, kidney, and vascular cells [21-26]. Animal viral pneumonia models provided indirect support showing improved outcome with losartan, including amelioration of lung injury, edema, and lung failure in a SARS-CoV infection model [26], and of lung injury, edema, and leukocyte infiltration in an influenza A H5N1 mouse model [27].

Some researchers raised concern early in the pandemic that ACEi/ARB might worsen COVID-19 outcome [28-29] because in some (not all) animal models, ACE2 expression may be upregulated by these drugs potentially increasing virus cell injury [30-37]. Human studies do not show increased plasma ACE2 levels with ACEi/ARB [38-40]. ARB have been hypothesized to stabilize with the ATR-1-ACE2 complex causing AT-1,7 generation leading to vasodilation and diminished inflammation [41]. These effects may be more important than ACE-2 inhibition/viral entry inhibition.

In sum, animal studies have shown potential for benefit with ACEi/ARB in viral pneumonias and inconsistent data suggesting potentially adverse increased ACE-2 expression.

### Retrospective/observational clinical trials

A Chinese retrospective study compared mortality in hypertensive hospitalized COVID-19 patients who were taking ACEi/ARB or were not [1]. Unadjusted and adjusted mortality was lower with ACEi/ARB. In another Chinese retrospective study in hypertensive hospitalized COVID-19 patients, progression to severe disease/mortality was similar in those taking vs. not taking ACEi/ARB [2]. An observational database of international hospitalized COVID-19 patients showed similar in-hospital mortality for ACEi/ARB use [3]. A NYU database review found no association for ACEi/ARB use and COVID-19 or severe COVID-19 [4]. An Italian case-control study found no association for ACEi/ARB use and COVID-19 [5].

In another hospitalized COVID-19 patient cohort, adjusted death/ICU transfer occurred less frequently with ACEi vs. non-ACEi use [6]. Danish national registry retrospective cohort and case-control studies found no significant associations for mortality and composite mortality/severe COVID-19 in patients taking vs. not taking ACEi/ARB [7]. An observational trial evaluating hydroxychloroquine/chloroquine found baseline ACEi (but not ARB) use to be associated with decreased mortality [3]. A Cleveland Clinic Health System retrospective cohort study reported no association for ACEi/ARB use and COVID-19 [8]. In a retrospective Stanford study of COVID-19 inpatients/outpatients, ACEi/ARB use was not associated with higher hospitalization, ICU risk, or death; hospitalization was lower [9].

In sum, these retrospective studies showed no safety signals for ACEi or ARB in COVID-19 [10] and possible benefit in some, supporting RCTs.

### Retrospective/observational clinical trials in influenza

A query of UK’s Clinical Practice Research Datalink showed lower influenza rates with ACEi/ARB use vs. non-use [42]. Influenza A relies on the ACE2 receptor for lung entry, like SARS-CoV-2.

### Prospective clinical trials

The BRACE CORONA trial compared discontinuation vs. continuation of ACEi/ARB already taken by 659 mild-moderate hospitalized COVID-19 patients in Brazil [11]. Median time from symptoms onset was 6 days. The trial found no differences in the primary outcome of days alive outside the hospital at day 30 (mean, 21.9 vs. 22.9 days). No significant differences in mortality and AEs were seen.

A second similar discontinuation vs. continuation trial of ARB already taken by 152 hospitalized COVID-19 patients was also published (REPLACE COVID) [12]. Median time from symptoms onset was 6.5-6.8 days. There was no difference in the study’s primary outcome, a global rank score including time-to-death, mechanical ventilation time, renal replacement, vasopressor time, and MOF. Mortality (13 vs. 15%), ICU admission/mechanical ventilation (18 vs. 21%), and AEs (36 vs. 39%) did not differ.

We are aware of five published ACEi/ACE in COVID-19 intervention trials (four inpatient, one outpatient).

The first, from University of Kansas, was a single-arm open-label trial assessing the ARB losartan in hospitalized COVID-19 patients requiring oxygen [13]. Thirty active treatment patients vs. 30 post-hoc external controls using propensity scores were compared, with the primary outcome being AE incidence. AE incidence (80 vs. 97%) and AE rates were lower with losartan (2.2 vs. 3.3); Poisson regression adjusted AE incidence ratio remained lower with losartan (0.69; 95% CI: 0.49-0.97); elevated creatinine AEs occurred in 30 vs. 23%, and elevated AST in 33 vs. 63%. No significant differences were found for death (1/30 vs. 3/30) or mechanical ventilation (2/17 vs. 5/17) (trends favored losartan); hospital and ICU LOS and days requiring oxygen or mechanical ventilation were similar. Study limitations included single-arm, nonrandomized, open-label design with external historical controls, and between-group imbalance. 34 of 347 screened patients were enrolled (10%).

The second, an open-label RCT compared the ARB telmisartan and SOC vs. SOC alone in two hospitalized COVID-19 patients in Argentina up to 4 days post symptoms onset (earlier than our trial) [14]. The primary outcome was CRP at days 5 and 8. In an interim analysis, 40 telmisartan/SOC vs. 38 SOC were compared. Mean CRP was significantly lower in telmisartan/SOC vs. SOC groups at days 5 (24.2 vs. 51.1 mg/L [p<0.05]) and 8 (9.0 vs. 41.6 mg/L [p<0.05]). Median time to discharge was shorter with telmisartan, 9 vs. 15 days (p=0.01) and 30-day mortality trend favored telmisartan (5.26 vs. 11.76%, p=0.41); there were no differences for ICU admission, mechanical ventilation, and a composite of ICU admission, mechanical ventilation, and death; fewer patients receiving telmisartan needed oxygen at day 15 (2/4 vs. 13/14, p<0.05). No telmisartan related AEs occurred and BP was similar. 82 of 185 screened patients were enrolled (56% [higher than our trial]).

The third, an open-label RCT compared the ARB losartan and SOC vs. SOC alone in 31 SHARP (San Diego) hospitalized patients with COVID-19 with mild hypoxia (requiring </= 3 L oxygen) [15]. The primary measurement was a composite of mechanical ventilation/death. No significant primary or secondary differences were found: the composite endpoint occurred in 1/16 vs. 1/15 losartan/SOC vs. SOC patients; mortality occurred in 1/16 vs. 1/15 patients; mean LOS was 9 vs. 10 days; hypotension occurred in 3/16 vs. 4/15. O2 requirements were similar.

The fourth, the first blinded placebo-controlled RCT, compared losartan to placebo in outpatients with mild COVID-19 conducted by University of Minnesota [17]. Of 117 enrolled patients, hospitalization (primary outcome) occurred in 3/58 (5.2%) vs. 1/59 (1.7%) of losartan vs. placebo patients – an insignificant absolute difference of -3.5% favoring placebo (95% CI -13.2, 4.8%; p=0.320). ICU admissions and viral loads were similar; there were no deaths. The rate of COSS <5 at day 15 was 7/55 (12.7%) with losartan and 2/54 (3.6%) with placebo, with an adjusted OR (age and gender) of -1.4 (95% CI -3.4, 0.2; p = 0.096), favoring placebo. AE rates were similar, 0.33 vs. 0.37, respectively; SAE rates were similar. No significant AKI, hypotension, or hyperkalemia occurred. This RCT was terminated early due to futility due low rates of events. 117 of 14,371 screened patients were enrolled (0.81%); 89.8% did not meet inclusion/exclusion criteria.

The fifth, also by University of Minnesota, was the second randomized placebo-controlled RCT, comparing losartan (N=101) and placebo (N=104) in hospitalized COVID-19 patients [18]. Losartan dosing was 50 mg twice daily for 10 days (higher than in our study). The primary outcome, the PaO2/FiO2 ratio at 7 days was similar [difference -24.8 (95% CI, -55.6 to 6.1; p=0.12)]. There were no differences in secondary outcomes including 90-day mortality (10.9 vs. 10.6%, respectively), LOS, oxygen and mechanical ventilation requirement at 10 days, or COSS. More losartan vs. placebo patients needed vasopressors (20 vs. 11%, p=0.08) and vasopressor-free days were lower with losartan (8.7 vs. 9.4, p=0.04); our study found numerically higher hypotension AE/SAE rates analogously. Overall AE and SAE rates were numerically higher with losartan (not significant); AKIs were more common with losartan (19.4 vs. 8.8%, p=0.04). This trial’s design and applicability was most like ours, including low enrollment:screening (5%). Notable differences include larger N, higher dosing, and lower mortality; similarities include lack of efficacy findings accompanied by numerically higher safety signals with losartan.

In sum, the ACEi/ARB continuation:discontinuation trials and small unblinded treatment trials in hospitalized COVID-19 patients showed equivalent or improved outcome. In contrast, both blinded RCTs showed lack of efficacy and adverse safety signals [17-18].

### Meta-analyses

A recent ACEi/ARB COVID-19 continuation:discontinuation meta-analysis (52 studies – 101,949 patients, 26,545 ACEi/ARB) found lower adjusted mortality and SAE rates with ACEi/ARB use [16].

Two pooled analyses of ACEi/ARB in COVID-19 are currently being completed and results should be forthcoming: (1) an IPD-based analysis of North American trials in hospitalized COVID-19 patients; (2) a larger aggregate data-based international meta-analysis by the International Society of Hypertension of outpatient and hospitalized COVID-19 patients [43]. COVID MED data are included in both.

### Guidelines, professional societies, and reviews

NIH guidelines, professional societies, and reviews recommend that ACEi/ARB, if already taken, should be continued but not started anew outside RCTs [10, 29-30, 44-49].

### Study strengths and limitations

The strengths of our study include its design (randomized, controlled, blinded), minimal primary outcome measurements missingness, and baseline balance. Limitations include small N, early termination, low enrollment rate making conclusions relevant to a small proportion of hospitalized COVID-19 patients (lowering external validity), and post-hoc SAP alterations.

### Conclusions

Our blinded and controlled RCT comparing losartan and control in hospitalized COVID-19 patients found no significant efficacy effect (COSS or mortality) which is not surprising given its small size, yet it found potential safety signals. We speculate that class adverse effects of ACEi/ARB may overcome theorized SARS-CoV-2-ACE-2 binding inhibition and other potential benefits making the overall benefit to risk ratio for these medications in COVID-19 null or negative. Given that results from our blinded RCT simulate those of the other two larger blinded RCTs to date, in effect a small third ‘negative’ RCT, unless forthcoming pooled analyses find conflicting results, ARBs should not be started *de novo* to treat COVID-19 outside of clinical trials.

## Supporting information

CONSORT checklist

ICF

EQUATOR checklist

## Data Availability

All data produced in the present study are available upon reasonable request to the authors.

## Acknowledgements

We thank our study patients for participating in this RCT during the difficult times of the COVID-19 pandemic. We thank Jessica Kumar, MD, and colleagues at New York State Department of Health, Albany, NY, for assistance with nasopharyngeal PCR processing and study drug supply.

## REFERENCES

1. Zhang P, Zhu L, Cai J, Lei F, Qin J-J, Xie J, et al. Association of Inpatient Use of Angiotensin Converting Enzyme Inhibitors and Angiotensin II Receptor Blockers with Mortality Among Patients With Hypertension Hospitalized with COVID-19. Circulation Research 2020;126:1671–1681.

2. Li J, Wang X, Chen J, Zhang H, Deng D. Association of Renin-Angiotensin System Inhibitors With Severity or Risk of Death in Patients With Hypertension Hospitalized for Coronavirus Disease 2019 (COVID-19) Infection in Wuhan, China. JAMA Cardiol 2020;5(7):825–830.

3. Mehra MR, Desai SS, Kuy SR, Henry TD, Patel AN. Cardiovascular Disease, Drug Therapy, and Mortality in Covid-19. N Engl J Med 2020;382:e102.

4. Reynolds HR, Adhikari S, Pulgarin C, Troxel AB, Iturrate E, Johnson SB, et al. Renin–Angiotensin– Aldosterone System Inhibitors and Risk of Covid-19. N Engl J Med 2020;382:2441–2448.

5. Mancia G, Rea F, Ludergnani M, Apolone G, Corrao G. Renin–Angiotensin–Aldosterone System Blockers and the Risk of Covid-19. N Engl J Med 2020;382:2431–2440.

6. Bean DM, Kraljevic Z, Searle T, Bendayan R, Kevin O′ G, Pickles A, et al. Angiotensin-converting enzyme inhibitors and angiotensin II receptor blockers are not associated with severe COVID-19 infection in a multi-site UK acute hospital trust. Eur J Heart Fail 2020;22(6):967–974.

7. Fosbol EL, Butt JH, Østergaard L, Andersson C, Selmer C, Kragholm K, et al. Association of Angiotensin-Converting Enzyme Inhibitor or Angiotensin Receptor Blocker Use With COVID-19 Diagnosis and Mortality. JAMA 2020;324(2):168–177.

8. Mehta N, Kalra A, Nowacki AS, Anjewierden S, Han Z, Bhat P, et al. Association of Use of Angiotensin-Converting Enzyme Inhibitors and Angiotensin II Receptor Blockers With Testing Positive for Coronavirus Disease 2019 (COVID-19). JAMA Cardiol 2020;5(9):1020–1026.

9. Rubin SJS, Falkson SR, Degner NR, Blish CA. Safety of ACE-I and ARB medications in COVID-19: A retrospective cohort study of inpatients and outpatients in California. J Clin Transl Sci 2020; doi: 10.1017/cts.2020.489.

10. Jarcho JA, Ingelfinger JR, Hamel MB, D’Agostino Sr RB, Harrington DP. Inhibitors of the Renin– Angiotensin–Aldosterone System and Covid-19. N Engl J Med 2020;382(25):2462–2464.

11. Lopes RD, Macedo AVS, de Barros E Silva PGM, Moll-Bernardes RJ, dos Santos TM, Mazzaet L, al. Effect of Discontinuing vs Continuing Angiotensin-Converting Enzyme Inhibitors and Angiotensin II Receptor Blockers on Days Alive and Out of the Hospital in Patients Admitted With COVID-19A Randomized Clinical Trial. JAMA 2021;325(3):254–264.

12. Cohen JB, Hanff TC, William P, Sweitzer N, Rosado-Santander NR, Medinaet C, al. Continuation versus discontinuation of renin–angiotensin system inhibitors in patients admitted to hospital with COVID-19: a prospective, randomised, open-label trial. Lancet Respir Med 2021;9(3):275–284.

13. Bengtson CD, Montgomery RN, Nazir U, Satterwhite L, Kim MD, Bah NC, et al. An Open Label Trial to Assess Safety of Losartan for Treating Worsening Respiratory Illness in COVID-19. F Front Med (Lausanne) 2021; doi: 10.3389/fmed.2021.630209.

14. Duarte M, Pelorosso F, Nicolosi LN, Salgado MV, Vetulli H, Aquieri A, et al. Telmisartan for treatment of Covid-19 patients: An open multicenter randomized clinical trial. EClinicalMedicine 2021; doi: 10.1016/j.eclinm.2021.100962.

15. Geriak M, Haddad F, Kullar R, Greenwood KL, Habib M, Habib C, et al. Randomized Prospective Open Label Study Shows No Impact on Clinical Outcome of Adding Losartan to Hospitalized COVID-19 Patients with Mild Hypoxemia. Infect Dis Ther 2021;10(3):1323–1330.

16. Baral R, Tsampasian V, Debski M, Moran B, Garg P, Clark A, et al. Association Between Renin-Angiotensin-Aldosterone System Inhibitors and Clinical Outcomes in Patients With COVID-19A Systematic Review and Meta-analysis. JAMA Netw Open 2021;4(3):e213594.

17. Puskarich MA, Cummins NW, Ingraham NE, Wacker DA, Reilkoff RA, Driver BE, et al. A multi-center phase II randomized clinical trial of losartan on symptomatic outpatients with COVID-19. EClinicalMedicine 2021; doi: 10.1016/j.eclinm.2021.100957.

18. Puskarich MA, Cummins NW, Ingraham NE, Wacker DA, Black LP, et al. A multi-center phase II randomized clinical trial of losartan on symptomatic outpatients with COVID-19. EClinicalMedicine. 37. 100957. 10.1016/j.eclinm.2021.100957.

19. Horby P, Mafham M, Linsell L, Bell JL, Staplin N, Emberson JR, et al. Effect of Hydroxychloroquine in Hospitalized Patients with Covid-19. The RECOVERY Collaborative Group. N Engl J Med 2020;383(21):2030–2040.

20. Cao B, Wang Y, Wen D, Liu W, Wang J, Fan G, et al. A Trial of Lopinavir–Ritonavir in Adults Hospitalized with Severe Covid-19. N Engl J Med 2020;382(19):1787–1799.

21. Wan Y, Shang J, Graham R, Baric RS, Li F. Receptor Recognition by the Novel Coronavirus from Wuhan: an Analysis Based on Decade-Long Structural Studies of SARS Coronavirus. J Virol 2020; doi: 10.1128/JVI.00127-20. Print 2020 Mar 17.

22. Letko M and Munster V, 2020. Functional assessment of cell entry and receptor usage for lineage B β-coronaviruses, including 2019-nCoV. Nat Microbiol 2020; https://doi.org/10.1038/s41564-020-0688-y.

23. Zhou P, Yang X-L, Wang X-G, Hu B, Zhang L, Zhang W, et al. A pneumonia outbreak associated with a new coronavirus of probable bat origin. Nature 2020;579:270–273 (2020).

24. Lu R, Zhao X, Li J, Niu P, Yang B, Wu H, et al. Genomic characterisation and epidemiology of 2019 novel coronavirus: implications for virus origins and receptor binding. Lancet 2020;395(10224):565–574.

25. Hoffmann M, Kleine-Weber H, Schroeder S, Krüger N, Herrler T, Erichsen S, et al. SARS-CoV-2 Cell Entry Depends on ACE2 and TMPRSS2 and Is Blocked by a Clinically Proven Protease Inhibitor. Cell 2020;181(2):271–280.

26. Kuba K, Imai Y, Rao S, Gao H, Guo F, Guanet B, et al. A crucial role of angiotensin converting enzyme 2 (ACE2) in SARS coronavirus–induced lung injury. Letter. Nat Med 2005;11:875–879.

27. Yan Y, Liu Q, Li N, Du JC, Li X, Li C, et al. Angiotensin II receptor blocker as a novel therapy in acute lung injury induced by avian influenza A H5N1 virus infection in mouse. LETTER TO THE EDITOR. Sci China Life Sci 2015;58(2):208–11.

28. Fang L, Karakiulakis G, Roth M. Are patients with hypertension and diabetes mellitus at increased risk for COVID-19 infection? Lancet Respir Med 2020; doi: 10.1016/S2213-2600(20)30116-8.

29. Vaduganathan M, Vardeny O, Michel T, McMurray JJV, Pfeffer MA, Solomon SD. Renin– Angiotensin–Aldosterone System Inhibitors in Patients with Covid-19. N Engl J Med 2020; 382:1653–1659.

30. Bozkurt B, Kovacs R, Harrington B. Joint HFSA/ACC/AHA Statement Addresses Concerns Re: Using RAAS Antagonists in COVID-19. J Card Fail 2020; doi: 10.1016/j.cardfail.2020.04.013.

31. Ferrario CM, Jessup J, Chappell MC, Averill DB, Brosnihan KB, Tallant EA, et al. Effect of Angiotensin-Converting Enzyme Inhibition and Angiotensin II Receptor Blockers on Cardiac Angiotensin-Converting Enzyme 2. Circulation 2005;111(20):2605–10.

32. Zheng Y, Ma Y-T, Zhang J-Y, Xie X. COVID-19 and the cardiovascular system. Nat Rev Cardiol 2020;17(5):259–260.

33. Ocaranza MP, Godoy I, Jalil JE, Varas M, Collantes P, Pinto M, et al. Enalapril Attenuates Downregulation of Angiotensin-Converting Enzyme 2 in the Late Phase of Ventricular Dysfunction in Myocardial Infarcted Rat. Hypertension 2006;48(4):572–8.

34. Ishiyama Y, Gallagher PE, Averill DB, Tallant EA, Brosnihan KB, Ferrario CM. Upregulation of Angiotensin-Converting Enzyme 2 After Myocardial Infarction by Blockade of Angiotensin II Receptors. Hypertension 2004;43(5):970–6.

35. Soler MJ, Ye M, Wysocki J, William J, Lloveras J, Batlle D. Localization of ACE2 in the Renal Vasculature: Amplification by Angiotensin II Type 1 Receptor Blockade Using Telmisartan. Am J Physiol Renal Physiol 2009;296(2):F398–405.

36. Burrell LM, Risvanis J, Kubota E, Dean RG, MacDonald PS, Lu S, et al. Myocardial infarction increases ACE2 expression in rat and humans. Eur Heart J 2005;26(4):369–75.

37. Burchill LJ, Velkoska E, Dean RG, Griggs K, Patel SK, Burrell LM. Combination renin-angiotensin system blockade and angiotensin-converting enzyme 2 in experimental myocardial infarction: implications for future therapeutic directions. Clin Sci (Lond) 2012;123(11):649–58.

38. Walters TE, Kalman JM, Patel SK, Mearns M, Velkoska E, Burrell LM. Converting enzyme 2 activity and human atrial fibrillation: increased plasma angiotensin converting enzyme 2 activity is associated with atrial fibrillation and more advanced left atrial structural remodeling. Europace 2017;19(8):1280–1287.

39. Ramchand J, Patel SK, Srivastava PM, Farouque O, Burrell LM. Elevated Plasma Angiotensin Converting Enzyme 2 Activity Is an Independent Predictor of Major Adverse Cardiac Events in Patients With Obstructive Coronary Artery Disease. PLoS One 2018; doi: 10.1371/journal.pone.0198144.

40. Furuhashi M, Moniwa N, Mita T, Fuseya T, Ishimura S, Ohno K, et al. Urinary angiotensin-converting enzyme 2 in hypertensive patients may be increased by olmesartan, an angiotensin II receptor blocker. Am J Hypertens 2015;28(1):15–21.

41. Li XC, Zhang J, Zhuo JL. The vasoprotective axes of the renin-angiotensin system: physiological relevance and therapeutic implications in cardiovascular, hypertensive and kidney diseases. Pharmacol Res 2017;125(Pt A):21-38.

42. Chung S-C, Providencia R, Sofat R. Association between Angiotensin Blockade and Incidence of Influenza in the United Kingdom. N Engl J Med 2020;383(4):397–400.

43. Gnanenthiran SR, Borghi C, Burger D, Charchar F, Poulter NR, Schlaich MP, et al. Prospective meta-analysis protocol on randomised trials of renin–angiotensin system inhibitors in patients with COVID-19: an initiative of the International Society of Hypertension. BMJ Open 2021; doi: 10.1136/bmjopen-2020-043625.

44. NIH COVID-19 Treatment Guidelines. (Updated August 4, 2021. Accessed 21 September 2021.) Available from https://www.covid19treatmentguidelines.nih.gov/therapies/concomitant-medications/.

45. de Simone G. Position Statement of the ESC Council on Hypertension on ACE-Inhibitors and Angiotensin Receptor Blockers. European Society of Cardiology. (13 March 2021; Accessed 1 October 2021.) Available from https://www.escardio.org/Councils/Council-on-Hypertension-(CHT)/News/position-statement-of-the-esc-council-on-hypertension-on-ace-inhibitors-and-ang.

46. Patel AB and Verma A. Nasal ACE2 Levels and COVID-19 in Children. JAMA 2020;323(23):2386– 2387.

47. Patel AB and Verma A. COVID-19 and Angiotensin-Converting Enzyme Inhibitors and Angiotensin Receptor Blockers: What Is the Evidence? JAMA 2020;323(18):1769–1770.

48. Lubel J and Garg M. Renin–Angiotensin–Aldosterone System Inhibitors in Covid-19. N Engl J Med 2020;382(24):e92.

49. Curfman G. Renin-Angiotensin-Aldosterone Inhibitors and Susceptibility to and Severity of COVID-19. JAMA 2020;324(2):177–178.

