## Supplementary material for "COVID MED – An Early Pandemic Randomized Clinical Trial of Losartan for Hospitalized COVID-19 Patients": ICF

**Informed Consent for Participation in a Research Study**

### Bassett Healthcare Network

Project No. 1581969

**Study title:** **C**omparison **O**f therapeutics for Hospitalized patients infected with SARS-Co**V**-2 **I**n a pragmatic a**D**aptive rando**M**iz**ED** clinical trial during the COVID-19 pandemic (COVID MED Trial)

**Investigator:** Daniel Freilich, MD

**Study Summary:** You are being asked to take part in this research project (called the COVID MED Trial) because you have tested positive for SARS-CoV-2 ("COVID-19") and are being treated for this infection in the hospital. The purpose of this study is to see if adding an investigational medication to usual treatment helps people get better faster, have less morbidity (fewer complications), and less mortality (better survival). Patients will be randomly assigned to get losartan (Cozaar, a high blood pressure medication) or placebo in addition to usual (standard) supportive care. Losartan is approved by the Food and Drug Administration (FDA) for other indications. Participation in this study will last about 60 days. Details of study "visits" are described below. The number of patients who will participate at Bassett Medical Center is unknown and will depend on how many patients are hospitalized here at Bassett. Other hospitals are expected to participate in this study and may include up to 2,000 patients, but it is not known exactly how many patients will be enrolled. Individual patients may or may not directly benefit from the study, but future patients may benefit if any of the investigational treatments are found to be more effective, allow patients to spend less time in the hospital, and improve morbidity and mortality. Participation in this research study is voluntary and will include only people who choose to take part. You can choose not to join the study and receive standard supportive treatment and discuss other potential options and approaches with your doctor for getting access to these and other investigational medications. Please read this consent form carefully and take your time making your decision. Please ask your doctor or research staff to discuss this consent form with you and explain any words or information that you do not understand.

**Background:** Patients with COVID-19 who need hospitalization can be ill for days to weeks, and can suffer life-threatening complications such as respiratory failure, sepsis, heart and heart rhythm problems, and death. Standard treatment includes supportive care, often with mechanical ventilation (breathing machine) in the Intensive Care Unit (ICU). Current recommendations also include the use of the medications, remdesivir and dexamethasone, for the treatment of some hospitalized patients with COVID-19. The COVID MED trial does not prohibit you from receiving any approved or recommended treatment, from participating in other trials, or from receiving other experimental treatments for COVID-19. Doctors have also been empirically (educated guessing) giving investigational medications that are encouraging in preclinical studies (in the laboratory and in animal models) and limited clinical human studies mostly for other indications. National and professional experts recommend making these experimental medications available to patients with COVID-19 under the umbrella of a scientifically sound clinical trial rather than giving these medications in other fashions. Therefore, we are conducting this clinical trial to test and compare an investigational medication that is FDA-approved for other indications and that shows potential for benefiting patients with COVID-19 (losartan) versus standard care alone (placebo). Previous versions of this study plan


Approved Date: July 21, 2020
The Mary Imogene Bassett Hospital IRB

included hydroxychloroquine and lopinavir/ritonavir treatment arms, but were halted due to new information from other studies that suggested that they were not effective in treating COVID–19.

**Goals:** The main goal of the COVID MED Trial is to assess whether adding the investigational medication (losartan) to standard care alone helps patients hospitalized with COVID-19 get better faster, be discharged faster, have less morbidity (fewer complications) and less mortality (fewer deaths). The study will also assess if it is safe for patients who are hospitalized with COVID-19 to be given this medication.

**Procedures to be Followed:** This is a placebo controlled, double blind study**.** If you agree to participate, this means you will then be randomly (like a flip of a coin) picked to get standard care PLUS either losartan or placebo (which is the same as standard care alone). To increase the chance you get the investigational medication, randomization will be in a 2:1 ratio, which means you will have a 2 in 3 (67%) chance of getting the investigational medication. You, your provider, and study investigators, will not know which group you have been assigned to, but every enrolled patient also gets standard treatment as per current guidelines; only the pharmacy and the unblinded study nurse in the Research office will know what medication you will get (he/she will not share this information unless there is an emergency). All medications, whether investigational or placebo, will be given once daily for 14 days if tolerated. The course of treatment may be shortened if the medication is not tolerated. The medication dose might vary depending on how healthy your kidneys are.

Study visits on Days 0, 1-7, 14, 30, and 60 will take place while you are in the hospital, on the phone, by video visit, and/or at an outpatient clinic.

Research staff will collect the following information from all study participants:

- Basic personal demographic information (age, sex, race, etc.)
- Relevant clinical information from interviews and the medical record (symptoms, vital signs, laboratory and imaging results, new diagnoses, other medical history, etc.).

You will be followed for up to 60 days via brief interviews by study staff (about 15-30 minutes each) and collecting clinical information during this time period – these will occur at 0 days (upon enrollment), then daily for 1 week, and then at 14, 30 and 60 days after you enroll in the study (eleven times in total). On some of these days, we will collect nasopharyngeal specimens (from the back of your nose) for SARS-CoV-2 PCR studies (to see if the virus is still present in your pharynx [back of throat]); when two of these tests are negative, we will stop doing them. We will try to do the interviews in person but telephone or video interviews will be acceptable if need be for logistics reasons.

****Due to a potential lack of PCR collection supplies, there may be times during the study when investigators will not be able to do these tests and they will be omitted from the study plan.****

**Blood testing:** We might also collect additional blood specimens (up to 15 ml - about 3 blood tubes) on these days BUT only if not already tested by your clinical provider AND you agree by choosing “I agree” below. You do not have to agree to the blood draws to participate in the study. These blood samples will be used to run safety laboratories (routine blood tests such as blood counts, chemistries, and inflammatory markers) if not already done by your provider as part of routine care. If you also agree below, we will also store samples for future unspecified research; if you do not agree to store your blood samples for future use, we will destroy them upon completion of the study. There will be


Approved Date: July 21, 2020
The Mary Imogene Bassett Hospital IRB

NO genetic studies. All results will be recorded using a de-identified study number. These results will not be available to you or your provider.

Please choose below:

- I **agree** to extra blood draws during this study: ________ (initials) <u>Choose one option below:</u>
  - I **also agree** to storing my blood samples for future studies: _______(initials)
  - <u>**But I do NOT agree**</u> to storing my blood samples for future studies: _______ (initials)
- I **do NOT agree** to extra blood draws during this study: ______ (initials)

---

- Please also **choose** whether you agree to **storage of nasopharyngeal** specimens for future studies: Yes _____, No ______ : _______ (initials)

Data collected in this research will be de-identified and may be used for future research or distributed to another investigator for future research without your additional consent.

**Potential Benefits:**

a. <u>Benefits to You</u>: You may benefit from this study if you receive the investigational medication and it is shown to help patients get better faster, leave the hospital earlier, and decrease morbidity (fewer complications) and mortality (fewer deaths), but no benefit is guaranteed.

b. <u>Potential Benefits to Society</u>: The information learned from this study may help improve the treatment of patients with COVID-19 in the future.

**Potential Risks:** All medications can cause side effects, sometimes mild and temporary, sometimes longer or more severe, and rarely life-threatening or causing death. Losartan is FDA approved for the treatment of hypertension and other indications. It is generally well tolerated, but unanticipated side effects (non-serious and serious) or drug interactions can occur. Note that these general comments about tolerability of the investigational medications being studied in this trial are based on experience in standard indications. There is limited safety data available for this medication in COVID-19 from controlled trials and it is possible that side effects mild, serious, and even life threatening, might be worse than is predicted. However in observational trials in COVID-19 (uncontrolled trials), losartan and drugs like it (known as ACE inhibitors and ARB's) have been reported to be possibly beneficial. There is also the possibility, that the investigational medication could worsen outcome more and even increase morbidity (complications) and mortality (deaths); this is unlikely but possible.

<u>**Losartan:**</u>
Losartan is used to treat high blood pressure. Losartan is also used in patients with heart failure, after heart attacks, and to protect kidneys in some diabetic patients. It is generally well tolerated, especially for short courses as occurs in this trial.


Approved Date: July 21, 2020
The Mary Imogene Bassett Hospital IRB

The most commonly reported side effects are:

- chest pain in 2-4%,
- low blood pressure in 4%,
- fatigue in 4%,
- myasthenia (a muscle/nerve problem) in at least 4%,
- dizziness in 3%,
- increased potassium or glucose each in at least 4%,
- diarrhea in at least 4%,
- urinary tract infection in at least 4%,
- back pain in 2 to at least 4%,
- upper respiratory infection in 8%,
- cough in 3%,
- nasal congestion in 2%,
- anemia in at least 4%,
- lack of energy in at least 4%,

The following side effects are possible, but are rare (less than 2%):

- atrial fibrillation
- stroke
- edema (swelling)
- palpitations
- passing out
- depression
- drowsiness
- headache
- migraine
- tingling in
- sleep disorder
- itching/photosensitivity/rash/hives
- abdominal pain
- constipation
- nausea and vomiting
- impotence
- high blood pressure
- joint or muscle pain
- ear ringing
- shortness of breath
- vertigo

Most of these side effects are transient and reversible but they can be serious. Other less common side effects, both non-serious and serious, including (usually temporary) acute kidney injury (AKI) or worsening of chronic kidney disease, have been seen and can occur. To reduce, risk patients known to very low blood pressure, significant acute or chronic kidney disease, and/or high potassium levels are excluded from the study.

There are potential benefits and risks of taking losartan in COVID-19 patients. Losartan may lessen the ability of COVID-19 virus to attach to lung cells (a potential benefit); but it is possible that it also increases the number of receptors in the lung to which the virus can attach (a potential risk). Several observational studies (uncontrolled) have shown showed lower or similar mortality in hospitalized COVID-19 patients in COVID-19 patients taking losartan versus those not taking one of them. Most professional guidelines recommend that COVID-19 patients generally continue to take losartan and similar medications if already taken for other indications. Researchers still can’t tell for sure whether this medication is helpful or not, so a placebo-controlled clinical trial such as this one is still needed

Nasopharyngeal swabs may be minimally uncomfortable but only briefly (20 seconds). Also, if you agree to extra blood testing (above), there will be momentary discomfort from the needle used to draw blood; sometimes, blood drawing leads to a bruise, fainting in some individuals, and rarely an infection.


Approved Date: July 21, 2020
The Mary Imogene Bassett Hospital IRB

**Women of Childbearing Potential:** The potential risk to a fetus from the study treatment in the setting of a COVID-19 infection is unknown. If you are a female of reproductive potential, you must not be pregnant at the beginning of this research or become pregnant during the study. A pregnancy test will be required before you begin the study. You must agree to take reasonable and necessary precautions against becoming pregnant during the period of the investigation. The investigator will discuss appropriate precautions with you. If at any point during the study you believe there is any possibility that you might be pregnant, you must notify the investigator immediately.

If important new findings come up during your participation in this study that could change your decision to be in this study, you will be given information about those findings as soon as possible.

A description of this study and a summary of the results will be available on http://www.ClinicalTrials.gov as required by U.S. Law. This website will not include information that can identify you. You can search this Web site at any time.

**Alternatives to Participation in this Research Study:** You may decide to not join this study and receive usual care and have opportunity to potentially get these or other investigational medications using alternative approaches (e.g., other clinical trials, compassionate use application, empiric usage, etc.); please discuss whether additional opportunities are available and possible with your doctor. Deciding not to participate in the study will not affect your access to care through the Bassett Healthcare Network.

**Financial Responsibility:** There should be no additional costs to you for participating in this study. Study medication or placebo and additional laboratory tests beyond those already ordered by your clinical provider will be paid for by the study. All other treatments and costs related to your hospitalization will be billed to you or your insurance per usual procedures.

In the event of physical injury resulting from research procedures, financial compensation is not available. Medical treatment is available through Bassett Healthcare Network at established charges. You would be responsible for any charges not covered by the research study or your own health care insurance. Further information may be obtained from the Office of Risk Management of Bassett Healthcare Network at (607) 547-6690.

**Compensation:** You will not be paid for participating in this study.

**Research Funding:** This study is being funded by a grant from the E. Donnall Thomas Resident Research Program and by internal support from the Bassett Research Institute.

**Confidentiality of Records and HIPAA Authorization:** While every effort will be made to keep your information private, this cannot be guaranteed. Other people may need to see the information. While they normally protect the privacy of the information, they may not be required to do so by law. Results of the research may be presented at meetings or in publications, but your name will not be used.

The federal Health Insurance Portability and Accountability Act (HIPAA) requires your permission to use your health information created or used as part of the research. Health information may include


Approved Date: July 21, 2020
The Mary Imogene Bassett Hospital IRB

your research record, related information from your medical records, results of laboratory tests, and both clinical and research observations made while taking part in the research, screening logs, and case report forms**.**

If you agree to participate, your medical records containing protected health information (individually identifiable information about you) may be accessed for conducting this research, fulfilling regulatory duties, provision of treatment, to determine research results, to monitor your health status, and to measure effects of the study drugs**.** It may be audited to make sure the researchers are following regulations, policies and study plans.

Representatives of the following groups may have access to your medical records and research related records for the purposes of conducting this research, reviewing the results and for regulatory duties:

- The investigators and staff coordinating this research;
- The Mary Imogene Bassett Hospital Institutional Review Board members and staff;
- The Department of Health and Human Services;
- Government agencies such as the Food and Drug Administration (FDA) for drug studies

If you decide to take part, your authorization for this study will not expire unless you cancel it. The information collected during your participation may be kept indefinitely. If you decide to withdraw your authorization, you must contact **both** parties listed below, in writing, and let them know that you are withdrawing authorization to use your protected health information:

Privacy Officer
Bassett Healthcare Network
Health Information Management
One Atwell Road
Cooperstown, NY 13326

Dr. Daniel Freilich
Bassett Healthcare Network
Department of Medicine
One Atwell Road
Cooperstown, NY 13326

If you cancel your authorization, you will also be removed from the study. However, standard medical care and any other benefits to which you are otherwise entitled will not be affected. Canceling your authorization only affects uses and sharing of information after the study investigator gets your written request. Information gathered before then may need to be used and given to others. For example, by Federal law, we must send study information to the FDA for drug and device studies it regulates. Information that may need to be reported to FDA cannot be removed from your research records.

The effective date for this authorization is the date that you sign the consent form.

Once information is disclosed under this authorization to someone who is not a health care provider, the information is no longer protected by the HIPAA federal privacy rule and could be disclosed to others by the recipient.

As stated in the section on Voluntary Participation below, you can also refuse to sign this consent/authorization and not be part of the study. You can also decide that you want to leave the study at any time without canceling the authorization. By signing this consent form, you give permission to use and/or share your health information as stated above.


Approved Date: July 21, 2020
The Mary Imogene Bassett Hospital IRB

**Right to Withdraw:** You have the right to withdraw from the study at any time, for any reason, with no negative effects on your care within the Bassett Healthcare Network. If you decide to withdraw, study drug will no longer be provided to you. You may discuss your other treatment options with your doctor.

The study doctor(s) may decide to withdraw you from the study, without your consent, if it is deemed to be in your best interest (for example if you are having a potential side effect from the study medicine) or if you are unable to follow the study procedures as described in this document.

**Right to Ask Questions**: You have the right to ask to any questions you may have about this research. If you have questions later or concerns related to this study, or if you believe you may have developed an injury that is related to this research, you should contact Dr. Freilich at 607-547-4586. If you have questions regarding your rights as a research subject, you may contact the Institutional Review Board Office at 607-547-3670.

**Voluntary Participation**: Participation in this research study is voluntary. You are free to withdraw from this study at any time. Your withdrawal from this study or your refusal to participate will in no way affect your continuing medical care or access to medical services at Bassett Healthcare.

**Subject's statement:**

I willingly agree to participate in this research study. I have reviewed the content of this consent form. I have had a chance to ask questions and have them answered to my complete satisfaction.
My signature below certifies that I consent to and give permission for my participation in this investigation. I have received a signed copy of this informed consent agreement.

| ___________________________________ | ____________ | ____________ |
|---|---|---|
| Subject's Signature | Date | Time |

___________________________________
Subject's Name (printed)

**Legally Authorized Representative (Signature):**

This is to certify that the person signing below consents to and gives permission for the participation of

________________________________in this research study.

| ___________________________________ | ____________ | ____________ |
|---|---|---|
| Signature of Subject's Representative (if applicable) | Date | Time |

| ___________________________________ | ___________________________________ |
|---|---|
| Representative's Name (printed) | Representative's relationship to subject |

***[Legal guardian/representative signature required adults unable to consent themselves. A legally authorized representative may need to provide consent for a subject who lacks the mental capacity to provide consent.]***


Approved Date: July 21, 2020
The Mary Imogene Bassett Hospital IRB

**Witness Signature (only if necessary*):***

I, the undersigned, have witnessed the presentation of the material contained in this consent form and observed that ___________________________ appeared to understand and willingly agreed to participate.

A witness was necessary because the subject was unable to read/sign because of physical disability or illiteracy or because the consent was administered via phone/video due to isolation precautions : ______________________________________________________________________________

______________________________________ _____________

Witness Signature (only if applicable) Date

_____________________________________

Witness Name (printed)

***[Witness signature is required for subjects who are unable to read the consent form but are competent to give consent*]**

**Person Obtaining Consent:**

I, the undersigned, have defined and explained the studies involved to the above subject.

______________________________________ _____________

Signature of Person Obtaining Consent Date

______________________________________

Name of person obtaining consent (printed)

***[Only those approved by the IRB to recruit and enroll subjects for this research may obtain informed consent].***



Approved Date: July 21, 2020
The Mary Imogene Bassett Hospital IRB
